# Integrative Harmonization of Phenotypic and Genomic Data Improves Bone Mineral Density Prediction in Multi-Study Osteoporosis Research

**DOI:** 10.1101/2025.05.12.25327471

**Authors:** Anqi Liu, Jianing Liu, Lang Wu, Qing Wu

**Author notes:** **Correspondence:** Qing Wu, MD, ScD, Department of Biomedical Informatics, College of Medicine, The Ohio State University, 250 Lincoln Tower, 1800 Cannon Drive, Columbus, OH 43210.

## Abstract

**Purpose:** Harmonizing osteoporosis-related data across multiple datasets is essential for improving the accuracy and generalizability of bone mineral density (BMD) assessments. This study developed a harmonization framework to standardize phenotypic and genomic variables across three major U.S. osteoporosis datasets: GDBF, GWAS, and NHANES.

**Methods:** We standardized key phenotypic variables (BMD, body mass index (BMI), age, sex, and race/ethnicity) using cohort-specific data dictionaries and applied multiple imputations by chained equations (MICE) to manage missing data. Genomic data were harmonized using principal component analysis (PCA)-based batch effect corrections. Residual regression methods were applied to standardize BMD values. The effectiveness of harmonization on BMD prediction was evaluated using generalized estimating equations (GEE) and mixed-effects models.

**Results:** Post-harmonization, inter-study variability in BMI was significantly reduced (Ω² = 0.0028), and BMD associations with covariates remained consistent across datasets. Harmonized models showed improved predictive performance, with explained variance in BMD increasing (R² = 0.14). PCA confirmed the effective alignment of genetic data, reducing batch effects and improving cross-study compatibility.

**Conclusion:** This study demonstrates the feasibility and effectiveness of harmonizing phenotypic and genomic data for osteoporosis research. The harmonization framework enhances BMD prediction accuracy, supports more inclusive osteoporosis risk assessment, and improves the integration of multi-cohort datasets for future research. These findings highlight the potential of data harmonization in advancing precision medicine for osteoporosis prevention and management.

**Mini-Abstract:** Harmonizing osteoporosis datasets improves BMD prediction accuracy and enhances risk assessment. This study developed a harmonization framework integrating phenotypic and genomic data across three major datasets. After harmonization, predictive model performance improved, enabling better osteoporosis risk stratification and advancing precision medicine for fracture prevention.

## Introduction

Osteoporosis, a chronic condition characterized by reduced bone mineral density (BMD) and increased fracture risk, affects approximately 18.3% of the global population, with 1 in 3 women and 1 in 5 men over 50 predicted to experience an osteoporotic fracture[1, 2]. Despite its prevalence and significant burden on public health, osteoporosis often remains underdiagnosed and undertreated, contributing to avoidable morbidity and mortality globally [3, 4].

Genetic studies, particularly genome-wide association studies (GWAS), have identified numerous single-nucleotide polymorphisms (SNPs) associated with BMD and fracture risk, shedding light on the genetic basis of osteoporosis[5–7]. However, existing research is limited by challenges in harmonizing genomic and phenotypic data across diverse studies[8, 9]. Variability in data collection methods, demographic representation, and the lack of standardized protocols for defining key variables such as BMD and body mass index (BMI) impede the ability to perform robust, large-scale analyses[10]. For example, inconsistencies in SNP imputation, differences in measurement techniques, and underrepresentation of minority populations have limited the generalizability of findings to non-European ancestry groups[11–14].

Harmonizing phenotypic and genomic data is critical to overcoming these barriers and advancing osteoporosis research. By aligning variable definitions, standardizing measurement units, and addressing demographic imbalances, harmonized datasets enable more comprehensive analyses of gene-environment interactions, improve statistical power, and foster equitable research outcomes.[15–18] However, Previous harmonization efforts often lacked comprehensive approaches to simultaneously standardizing both phenotypic and genomic variables, limiting their generalizability and practical applicability. Significant obstacles remain, including biases introduced by missing data, measurement instrument incompatibility, and varying inclusion criteria across datasets.[19, 20].

This study aimed to address these challenges by implementing a harmonization framework across multiple major U.S. osteoporosis datasets. The framework focuses on:

1. Standardizing phenotypic variables such as BMD, BMI, and demographic factors across datasets.
2. Applying robust statistical techniques, including mixed-effects models and generalized estimating equations (GEE), to account for inter-study variability.
3. Harmonizing genomic data through stringent quality control and imputation protocols to enable integration across datasets.

By providing a harmonized dataset that integrates both phenotypic and genomic data, we aim to establish a foundation for harmonized large-scale investigations into the phenotypic and genetic determinants of osteoporosis. The insights derived from this harmonization framework have the potential to enhance the understanding of BMD variation across diverse populations, advancing the field toward more inclusive and effective strategies for osteoporosis prevention and management.

## Methods Section

### 2.1 Study Design and Data Sources

This study harmonized data from three major U.S. osteoporosis studies/datasets: the Genetic Determinants of Bone Fragility (GDBF) study, the Genome-Wide Association Study (GWAS) for Osteoporosis, and the National Health and Nutrition Examination Survey (NHANES). These datasets were selected because they include key bone health measures, such as BMD, and possess comprehensive phenotypic variables and genomic data for osteoporosis research, enabling a multi-dataset evaluation of phenotype harmonization.

The GDBF study is a cross-sectional investigation assessing genetic factors influencing peak BMD in premenopausal women[21]. The GWAS study is a longitudinal cohort designed to explore genome-wide associations with osteoporosis, emphasizing genetic markers relevant to bone health[22]. NHANES is a nationally representative, cross-sectional survey conducted by the National Center for Health Statistics, providing data on BMD and other health parameters[23]. This study specifically utilized data from the 2009–2010, 2013–2014, and 2017– 2020 cycles[24]. The overall harmonization process, including both phenotypic and genomic data, is summarized in **Figure 1**, which illustrates the multi-step approach used in this study, including multi-study phenotype harmonization and BMD-specific harmonization.

**Figure 1.**
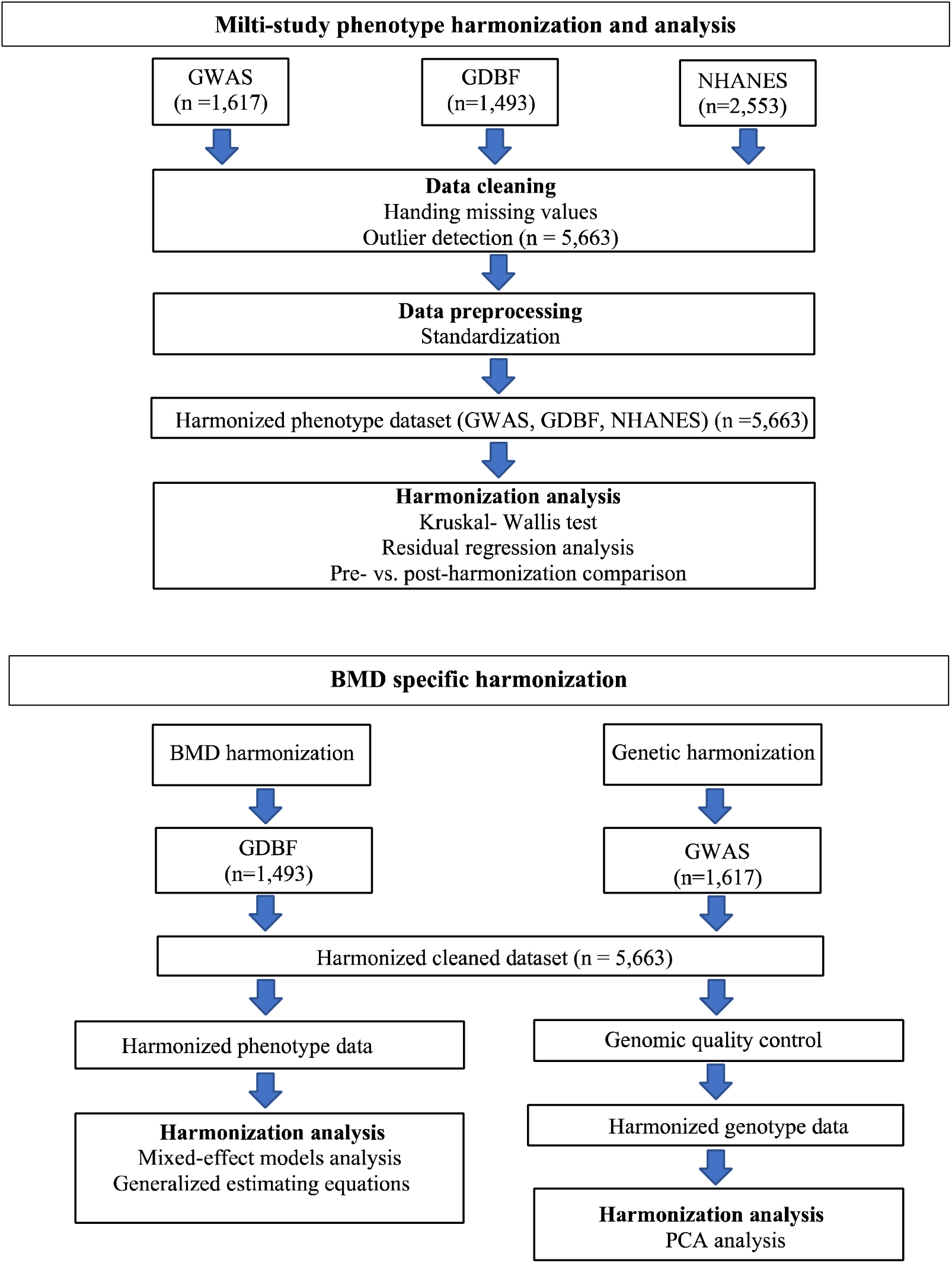
Harmonization Pipeline for Phenotypic and Genomic Data. Phenotypic harmonization involved data cleaning (handling missing values and outlier detection), followed by standardization and statistical analyses (Kruskal-Wallis test and residual regression) to ensure consistency across cohorts. Genomic harmonization of GWAS and GDBF datasets included rigorous quality control, imputation, and batch-effect correction, facilitating accurate integration and analysis.

Data were obtained from both controlled-access and publicly available repositories. The GDBF and GWAS datasets were accessed through the Database of Genotypes and Phenotypes (dbGaP). (dbGaP Study Accession: phs000390.v1.p1, phs000138.v2.p1). The NHANES was obtained from the National Center for Health Statistics (NCHS). Ethical approval was obtained from the relevant institutional review board (IRB #2025E0335).

#### Phenotypic Harmonization

In this study, bone mineral density (BMD) specifically refers to total hip BMD measured by dual-energy X-ray absorptiometry (DXA) and expressed in grams per square centimeter (g/cm²). BMD values in the GDBF cohort were pre-adjusted in dbGaP using residuals derived from linear regression models adjusting for baseline age and weight. To align methodologically with the GDBF adjustments, we applied the same residual regression approach separately within the GWAS cohort, replacing original BMD values with residuals computed from models adjusting for baseline age and weight. NHANES lacked hip BMD measurements and were therefore included solely for demographic comparisons.

Other key phenotypic variables—including BMI, age, sex, and race/ethnicity—were standardized using a structured harmonization framework. Variable definitions were consistently aligned across datasets through cohort-specific data dictionaries. BMI values were uniformly recalculated from height and weight measurements. Additionally, racial and ethnic categories were consolidated into four standardized groups: Black, Hispanic, White, and Other.

##### Handling of Missing Data and Imputation

Multiple imputations by chained equations (MICE) were applied separately within each cohort, assuming missing-at-random (MAR)[25, 26]. The imputation model included age, sex, BMI, race, and BMD to ensure unbiased estimates. Convergence diagnostics were performed, and post-imputation validation confirmed that the imputed values were closely aligned with observed data distributions.

### 2.3. Genomic Harmonization

Genomic data from GWAS and GDBF studies underwent rigorous quality control and harmonization.

#### Genotyping and Imputation

Genotyping was conducted using Illumina and Affymetrix arrays[27], followed by genotype imputation via the Haplotype Reference Consortium (HRC) panel using the Michigan Imputation Server[28]. Quality control measures included the removal of SNPs with a minor allele frequency (MAF) <0.01, call rate <99%, or Hardy-Weinberg equilibrium (HWE) p-value < 1e-5.

#### Population Structure and Batch Effect Correction

Principal component analysis (PCA) was used to evaluate population structure and detect batch effects[29, 30]. Scatter plots of PC1 vs. PC2 confirmed genetic similarity between the GWAS and GDBF datasets. Study-specific genomic batch effects were assessed and corrected, ensuring consistency across datasets. Quality control, PCA, and batch-effect corrections were performed using PLINK 2.0 and R statistical software (version 4.3.2), ensuring transparency and reproducibility.

### 2.4 Statistical Analysis

A combination of statistical approaches was applied to evaluate BMD associations and harmonization effectiveness.

#### Harmonization Effectiveness Assessment

Harmonization effectiveness was assessed by comparing the pre- and post-harmonization distributions of key phenotypic variables using Analysis of Variance (ANOVA) and Kruskal-Wallis tests. Residual regression analyses were performed to reduce inter-study variability, adjusting for age, sex, and race. Effect sizes (Omega squared, Ω²) were calculated to quantify the proportion of variance explained by study differences after harmonization[31].

#### BMD Association Analysis

BMD was modeled as a function of age, sex, BMI, and race using Generalized Estimating Equations (GEE)[32] for harmonized data, which accounted for the within-study correlation. Mixed-effects models were implemented[33], incorporating the study cohort as a random effect to capture inter-data variability. Additionally, separate linear regression models were applied to pre-harmonized GWAS and GDBF datasets to compare association estimates before and after harmonization.

#### Model Fit and Validation

Model performance was evaluated by comparing estimates across different statistical approaches. Adjusted R² (for linear regression), Pseudo R² (for mixed-effects models), and Quasi R² (for generalized estimating equations) were used to assess predictive accuracy, providing insights into model selection and comparative effectiveness.

### 2.5 Data Availability

The harmonized datasets and statistical analysis code are available upon reasonable request, subject to data use agreements (Accession: phs000138.v2. p1, phs000390.v1. p1). NHANES data used in this study are publicly available from the National Center for Health Statistics (NCHS) website (https://www.cdc.gov/nchs/nhanes/index.html).

## Results

### 3.1 Dataset Characteristics

The final harmonized dataset included 5,663 participants from three major datasets: GDBF (n=1,493), GWAS (n=1,617), and NHANES (n=2,553). **Table 1** presents the baseline characteristics before and after harmonization. Before harmonization, significant differences were observed across the datasets in age, BMI, sex, and racial composition (ANOVA, *p*<0.01). The mean BMI varied across datasets, with GDBF participants having a lower mean BMI (26.2 kg/m²) compared with NHANES (28.2 kg/m²) and GWAS (26.8 kg/m²). Similarly, the racial composition was notably different, with NHANES containing a more diverse population (39.5% White, 29.7% Hispanic, 20.6% Black, and 10.1% Other), whereas GWAS and GDBF were predominantly White (100% and 99.6%, respectively). The GDBF data consisted of younger participants (mean age: 32.7 years), whereas GWAS and NHANES participants had higher mean ages of 48.1 and 48.8 years, respectively.

**Table 1.** Participant Characteristics of the NHANES, GWAS, GDBF datasets and post-harmonization dataset.

| Characteristic | GDBF<br>(N = 1493) | GWAS<br>(N =1617) | NHANES<br>(N =2553) | Post-Harmonization<br>(N = 5663) | P-value** |
| --- | --- | --- | --- | --- | --- |
| Age(years) | 32.7(7.2) | 48.1(13.5) | 48.8(16.8) | 44.4(15.6) | <0.01 |
| Gender, n (%) |  |  |  |  | <0.01 |
| Female | 1493(100%) | 1209(74.8%) | 1288(50.5%) | 3990(70.5%) |  |
| Male |  | 408(25.2%) | 1265(49.5%) | 1673(29.5%) |  |
| Race, n (%) |  |  |  |  | <0.01 |
| Black |  |  | 526(20.6%) | 526(9.3%) |  |
| Hispanic | 6(0.4%) |  | 759(29.7%) | 765(13.5%) |  |
| White | 1487(99.6%) | 1617(100%) | 1009(39.5%) | 4113(72.6%) |  |
| Other |  |  | 259(10.1%) | 259(4.6%) |  |
| BMI (kg/m<sup>2</sup>) | 26.2(6.0) | 26.8(5.8) | 28.2(5.8) | 27.3(5.9) | <0.01 |

*BMI = body mass index*
\*\* *P*-values were calculated using ANOVA for continuous variables (Age and BMI) and Chi-square tests for categorical variables (Gender and Race) to assess differences across cohorts.

### 3.2 Missing Data Patterns and Imputation Outcomes

The extent of missing data varies across datasets, with GDBF exhibiting the highest proportion of missing BMD and BMI values. We found that up to 25.5% of BMD variables and 0.67% of BMI variables were missing in the GDBF study. In the NHANES study, 0.27% of BMI variables were missing. The GWAS study exhibited relatively complete phenotypic data, although additional quality control was required for some genomic markers due to low call rates.

Multiple imputations via the MICE approach were applied separately within each dataset. Post-imputation diagnostics confirmed that the imputed values closely matched the observed data distributions. Density plots comparing pre- and post-imputation distributions validated the imputation model, ensuring bias introduced by missing data to be minimized (**SFigure 2**).

**Figure 2.**
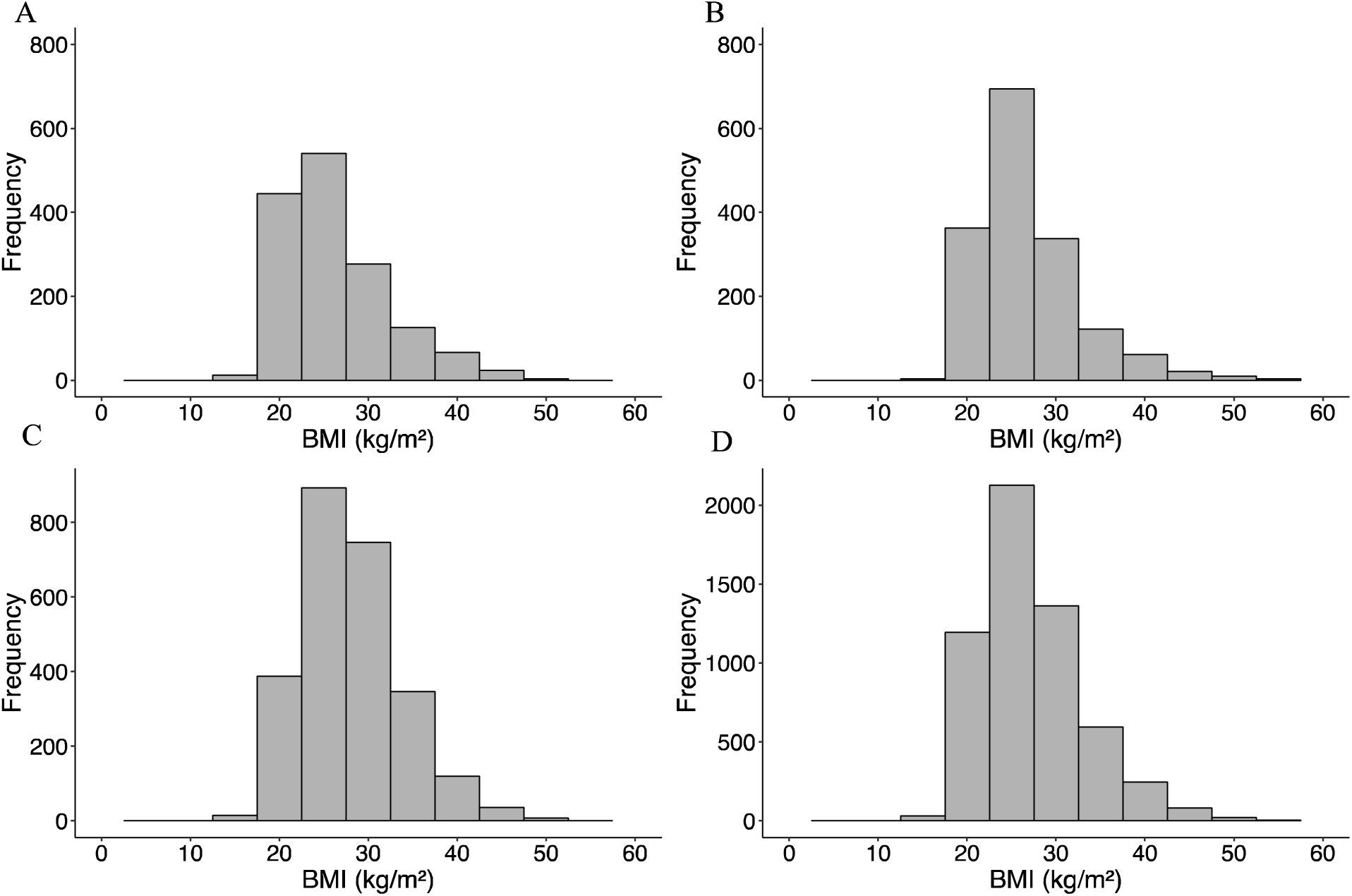
Comparison of Pre- and Post-Harmonization Distributions of BMI Across Datasets. (A) GDBF study, (B) GWAS study, and (C) NHANES cohort before harmonization; (D) Harmonized data across all cohorts, illustrating improved alignment.

### 3.3 Assessment of Harmonization Effectiveness

The differences in BMI across the GDBF, NHANES, and GWAS datasets were statistically significant, as indicated by the Kruskal-Wallis test (*p* <0.05 η² = 0.032), highlighting notable heterogeneities across study populations. **Figure 2** compares the pre- and post-harmonization distributions of BMI across the three datasets.

Before harmonization, the BMI distribution showed notable differences, with the GDBF cohort having a more skewed distribution towards lower BMI values compared with the NHANES and GWAS datasets. Post-harmonization, the BMI distributions across the three datasets became more aligned, reflecting the effectiveness of the harmonization process.

Post-harmonization adjustments for age, sex, and race significantly reduced inter-cohort variability, particularly in BMI distributions, as illustrated in **Figure 3**. The residual regression analyses confirmed that BMI differences across datasets decreased after harmonization. The calculated Omega squared (Ω² = 0.0028) indicated a meaningful reduction in the variance attributable to study differences, further supporting the success of the harmonization process. The overall BMI distribution became more aligned across datasets, demonstrating the efficacy of the harmonization process. The final datasets were well-balanced, enabling robust downstream analyses. The racial composition across participants after harmonization demonstrates a more balanced distribution across racial groups (**SFigure 1**).

**Figure 3.**
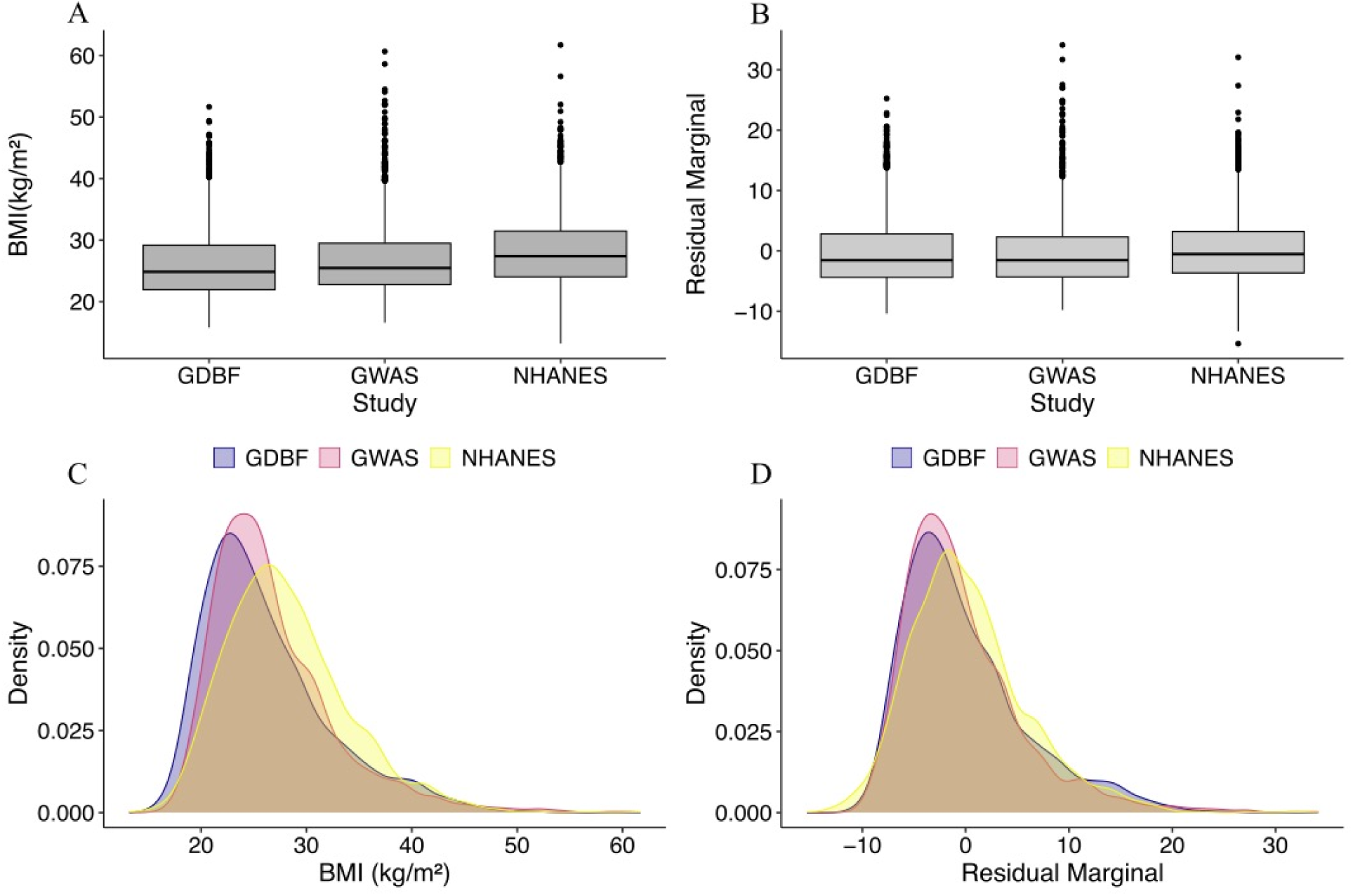
BMI Distributions Before and After Harmonization Adjustments. A) Boxplots illustrating BMI values across datasets before adjustment. (B) Boxplots of BMI residuals after adjustment for age, sex, and race. (C) Density plots of BMI values before adjustment, stratified by dataset. (D) Density plots of BMI residuals after adjustment for age, sex, and race, showing improved alignment across datasets.

**STable 1** provides further evidence of the success of the harmonization process, highlighting effect sizes and 95% confidence intervals (CI) for age and BMI across the GWAS, GDBF, and NHANES datasets before and after phenotype harmonization. Post-harmonization values were more consistent across datasets, demonstrating that cohort differences were minimized while retaining essential phenotypic information.

### 3.4 BMD Association Analysis and Model Performance

The associations between BMD and key covariates were evaluated using Generalized Estimating Equations (GEE) and Mixed-Effects Models. As detailed in **STable 2**, race and sex remained significant predictors for the GEE model.

After harmonization, the adjusted R² values increased from −0.07% (GDBF) and 0.002% (GWAS) to 0.14% (harmonized dataset), reflecting improved model performance and reduced inter-study variability (**Figure 4**). Although this increase was relatively modest, it is important for enabling robust comparative analyses and future pooled studies. The consistency of model estimates across different statistical approaches further confirms that the harmonization process effectively preserved essential phenotype-genotype relationships without introducing bias.

**Figure 4.**
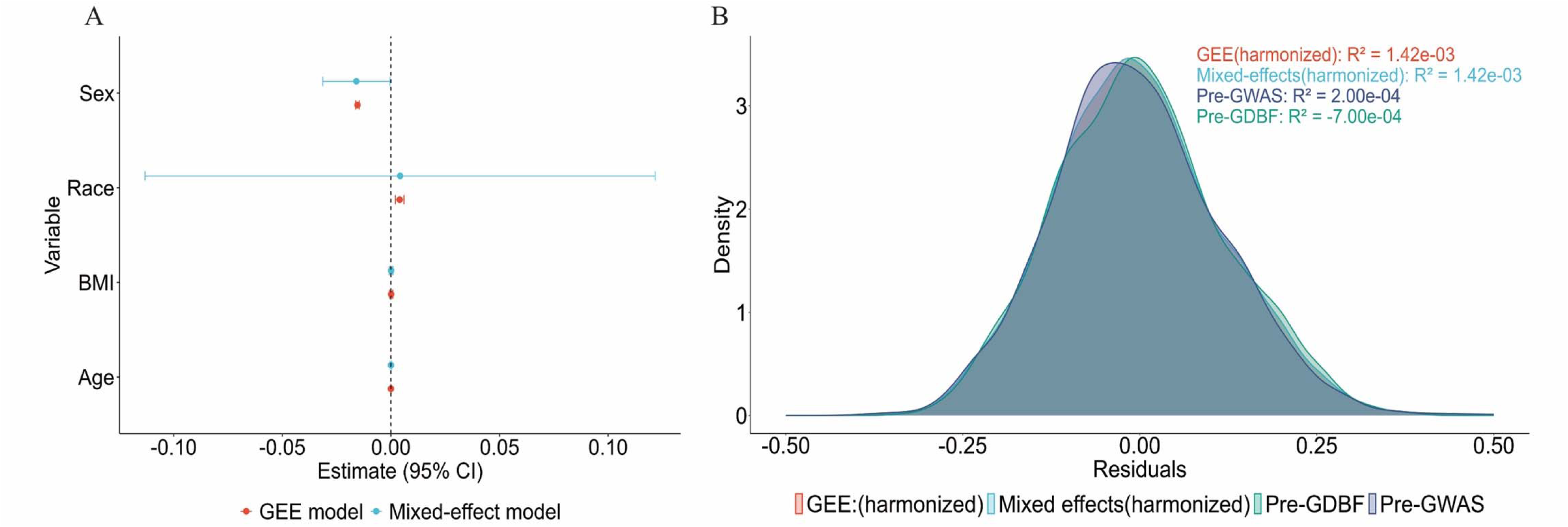
Comparison of Model Estimates and Residual Distributions for BMD Prediction. (A) Coefficient estimates with 95% confidence intervals for generalized estimating equations (GEE, red) and mixed-effects models (blue), assessing predictors (sex, race, BMI, and age) of BMD. (B) Density plots of residuals from pre- and post-harmonization models, showing consistent variance explained (R² values) by harmonized GEE and mixed-effects models across datasets

### 3.5 Population Structure and Genomic Harmonization Outcomes

Principal component analysis (PCA) was conducted to assess genomic harmonization between the GWAS and GDBF datasets. These analyses confirmed that no systematic biases remained after harmonization. The scatter plot of PC1 and PC2 (**Figure 5**) illustrates substantial overlaps between the two datasets, confirming that the genomic harmonization process effectively reduced batch effects. No significant clustering by cohort was observed, suggesting that population stratification was well-controlled in subsequent analyses.

**Figure 5.**
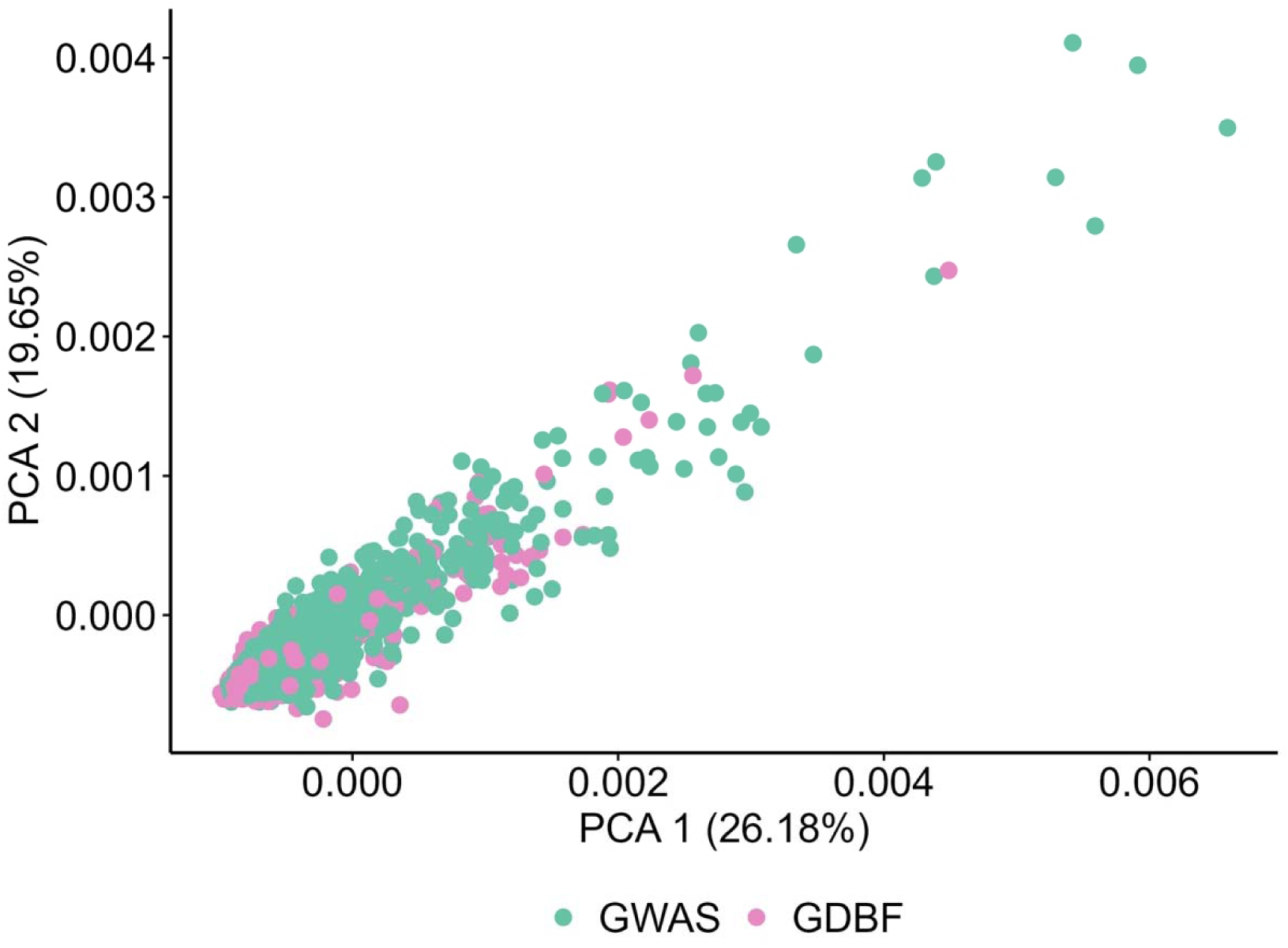
PCA Analysis of Genomic Harmonization. Scatter plot of the first two principal components (PC1: 26.18%; PC2: 19.65%), illustrating substantial overlaps between GDBF and GWAS datasets, indicating effective reduction of batch effects and successful genomic harmonization.

## Discussion

This study implemented a robust framework for harmonizing phenotypic and genomic data across three major U.S. osteoporosis datasets: GDBF, GWAS, and NHANES. The harmonized dataset addressed key challenges, including variability in phenotypic definitions, demographic imbalances, and missing data, enabling comprehensive analyses of BMD and its determinants[34]. Key findings include the successful harmonization of BMI, age, and race, reduction of inter-study variability, and validation of known associations between BMD and predictors such as age, BMI, and race.

The harmonization framework advanced previous approaches by comprehensively addressing inconsistencies in variable definitions and implementing robust statistical techniques, including multiple imputation and residual regression analysis[25, 35]. This approach ensured that the harmonized dataset was suitable for pooled analyses, while including diverse populations, particularly from NHANES, improved the generalizability of findings. Furthermore, genomic data were harmonized using stringent quality control and imputation protocols, enabling reliable genotype-phenotype association analyses.

This study builds upon and extends prior research in several key areas. Unlike previous studies[36–40] that often rely on single-cohort datasets, this work leveraged multiple datasets to enhance statistical power and representativeness. While genomic data integration has been extensively explored in other disease areas, its application to osteoporosis research remains relatively limited, primarily due to constraints in sample size[41, 42]. By harmonizing genomic and phenotypic data, we established a critical resource for investigating BMD and fracture risk across diverse populations. Additionally, the inclusion of NHANES improved demographic diversity compared with prior studies, which have predominantly focused on populations of European ancestry [43, 44].

The findings have significant implications for both clinical practice and research. The harmonized dataset lays the groundwork for developing more equitable and accurate fracture risk prediction models, which can account for population-specific differences, addressing long-standing disparities in osteoporosis diagnosis and treatment[45, 46]. Integrating genomic data also provides opportunities to explore gene-environment interactions, which may reveal new biological pathways underlying BMD and fracture risk [47]. By creating a comprehensive data harmonization framework, this study provides a valuable methodological resource that can facilitate future research, including meta-analyses and replication studies.

This study has several limitations. The study results generated from cross-sectional data may not be fully applicable to cohort data with phenotypic and genomic factors. Although multiple imputations effectively addressed missing data, some variables, such as alcohol consumption, were inconsistently recorded across datasets, potentially limiting the comprehensiveness of the analyses. Additionally, while NHANES improved demographic diversity, the overrepresentation of White participants in the GDBF and GWAS datasets highlights the need for further inclusion of underrepresented populations in future research. Furthermore, while harmonization aimed to enhance model performance by reducing variability and aligning definitions across datasets, the observed improvement in predictive accuracy was relatively modest. Several factors could potentially contribute to this limited increase, including prior adjustments for key confounders (age and weight) during harmonization and the inherently complex nature of osteoporosis risk.

Future directions include expanding the harmonization framework to longitudinal datasets and providing insights into the dynamic relationships between genomic factors and BMD over time. Efforts to include more racially and ethnically diverse populations are critical for ensuring the generalizability and equity of osteoporosis research. Additionally, incorporating other omics data, such as transcriptomics, methylomics, proteomics, and metabolomics, could further help to elucidate the molecular mechanisms underlying osteoporosis and related conditions[48, 49].

This study provides a robust and effective framework for harmonizing phenotypic and genomic data, facilitating the integration of multiple large-scale datasets. The harmonized data enhances the accuracy and generalizability of assessments, offering potential to develop more precise and equitable fracture risk prediction tools. By addressing methodological challenges such as inconsistent variable definitions, missing data, and demographic imbalances, this work establishes a foundation for future precision medicine research in osteoporosis, particularly for populations historically underrepresented in genomic studies.

The availability of these harmonized datasets presents significant opportunities for advancing osteoporosis research, enabling more precise identification of genetic and environmental determinants of bone health. Future studies can leverage this approach to investigate diverse populations, develop targeted preventive strategies, and ultimately reduce disparities in osteoporosis diagnosis and treatment.

## Data Sharing

The data/analyses presented in the current publication are based on study data downloaded from the dbGaP website under <u>phs000138.v2. p1</u> and <u>phs000390.v1. p1</u>, following the corresponding data use agreement. The NHANES data used in this study are publicly available from the National Center for Health Statistics (NCHS) at https://www.cdc.gov/nchs/nhanes/index.html.

## Disclosures of conflicts of interest

Anqi Liu, Jianing Liu, and Qing Wu declare that they have no conflict of interest. L.W. provided consulting services to Pupil Bio Inc., Techspert, and Galiher DeRobertis & Waxman LLP, and reviewed manuscripts for Gastroenterology Report, activities unrelated to this study, and received an honorarium.

## Funding

This study was supported by the National Institute on Minority Health and Health Disparities (R21MD013681, awarded to Dr. Q. Wu) and the National Institute on Aging (R01AG080017, awarded to Dr. Q. Wu).

## Role of the Funder/Sponsor

The funding sources had no role in the design and conduct of the study; collection, management, analysis, or interpretation of the data; preparation, review, or approval of the manuscript; or the decision to submit the manuscript for publication.

## Authors’ Contributions

Conceptualization: Qing Wu.

Data curation: Anqi Liu.

Formal analysis: Anqi Liu.

Funding acquisition: Qing Wu.

Investigation: Qing Wu, Anqi Liu.

Methodology: Anqi Liu, Qing Wu, Jianing Liu, Lang Wu

Project administration: Qing Wu.

Resources: Qing Wu.

Software: Anqi Liu.

Supervision: Qing Wu.

Validation: Anqi Liu.

Visualization: Anqi Liu.

Writing –original draft: Anqi Liu, Jianing Liu, Qing Wu.

Writing –review & editing: Qing Wu, Jianing Liu, Anqi Liu, Lang Wu.

## Notes

### Competing Interest Statement

The authors have declared no competing interest.

