## Supplementary Tables and Figures for "Integrative Harmonization of Phenotypic and Genomic Data Improves Bone Mineral Density Prediction in Multi-Study Osteoporosis Research"

**Supplementary Table 1**. Effect sizes and 95% confidence intervals (CI) for age and BMI across GWAS, GDBF, and NHANES datasets before and after phenotype harmonization.

| Characteristic | Post-Harmonization | GWAS (95% CI) | GDBF (95% CI) | NHANES (95% CI) |
| --- | --- | --- | --- | --- |
| Age | Reference | 0.25(0.19, 0.30) | -0.82 (-0.88, -0.76) | 0.28(0.23, 0.33) |
| BMI (kg/m²) | Reference | -0.08 (-0.13, -0.02) | -0.18 (-0.23, -0.12) | 0.15 (0.11, 0.20) |

Post-harmonization values are used as the reference dataset. Effect sizes were calculated using the standardized mean difference (SMD) method.

**Supplementary Table 2.** The estimated effects of key predictors on bone mineral density (BMD) using four different modeling approaches.

| Predictor | Model 1: GEE (Harmonized Data) | | Model 2: Mixed-Effects Model (Harmonized Data) | | Model 3: Linear Regression (GWAS Study) | | Model 4: Linear Regression (GDBF Study) | |
| --- | --- | --- | --- | --- | --- | --- | --- | --- |
|  | Estimate (95% CI) | P | Estimate (95% CI) | P | Estimate  (95% CI) | P | Estimate  (95% CI) | P |
| Age | 0.00 (0.00,0.00) | 0.83 | 0.00 (0.00,0.00) | 0.91 | 0.00  (0.00, 0.00) | 0.82 | 0.00  (0.00, 0.00) | 0.92 |
| Sex | -0.02  (-0.02, -0.02) | **<0.01** | -0.02  (-0.03, 0.00) | **0.04** | -0.02  (0.04,0.00) | 0.08 | - | - |
| Race | 0.00 (-0.02, 0.01) | **<0.01** | 0.00  (-0.11, 0.12) | 0.94 | - | - | 0.00  (-0.10, 0.10) | 0.97 |
| BMI (kg/m²) | 0.00 (0.00, 0.00) | 0.81 | 0.00  (0.00, 0.00) | 0.83 | 0.00  (0.00, 0.00) | 0.49 | 0.00  (0.00, 0.00) | 0.17 |

The Generalized Estimating Equations (GEE) model and the mixed-effects model were applied to the harmonized dataset, which integrates GWAS and GDBF data, while separate linear regression models were used for the pre-harmonized GWAS and GDBF datasets. Estimates are reported with 95% confidence intervals (CI), and corresponding p-values. Model 1: GEE model applied to harmonized data. Model 2: Mixed-effects model applied to harmonized data. Model 3: A linear regression model was applied to the pre-harmonized GWAS dataset. Race was not included as the GWAS study comprises only white population. Model 4: A linear regression model was applied to the pre-harmonized GDBF dataset. Sex was not included in the model since the GDBF study comprises only female participants.

**Supplementary Fig 1.** Proportion of racial groups among participants based on the harmonized "race" variable

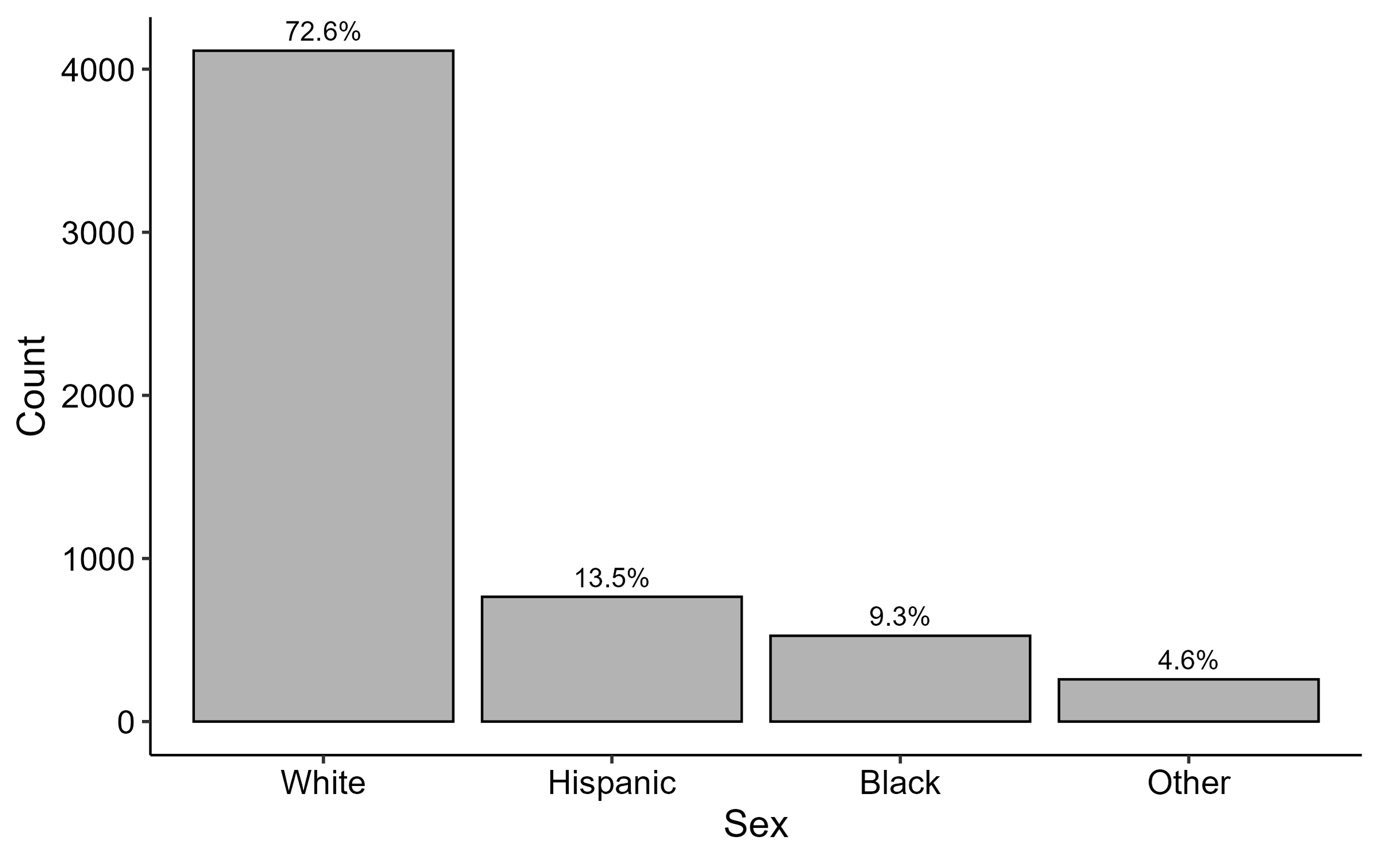

**Supplementary Fig 2.** Density plots comparing pre- and post-imputation distributions to validate the imputation model for each study cohort

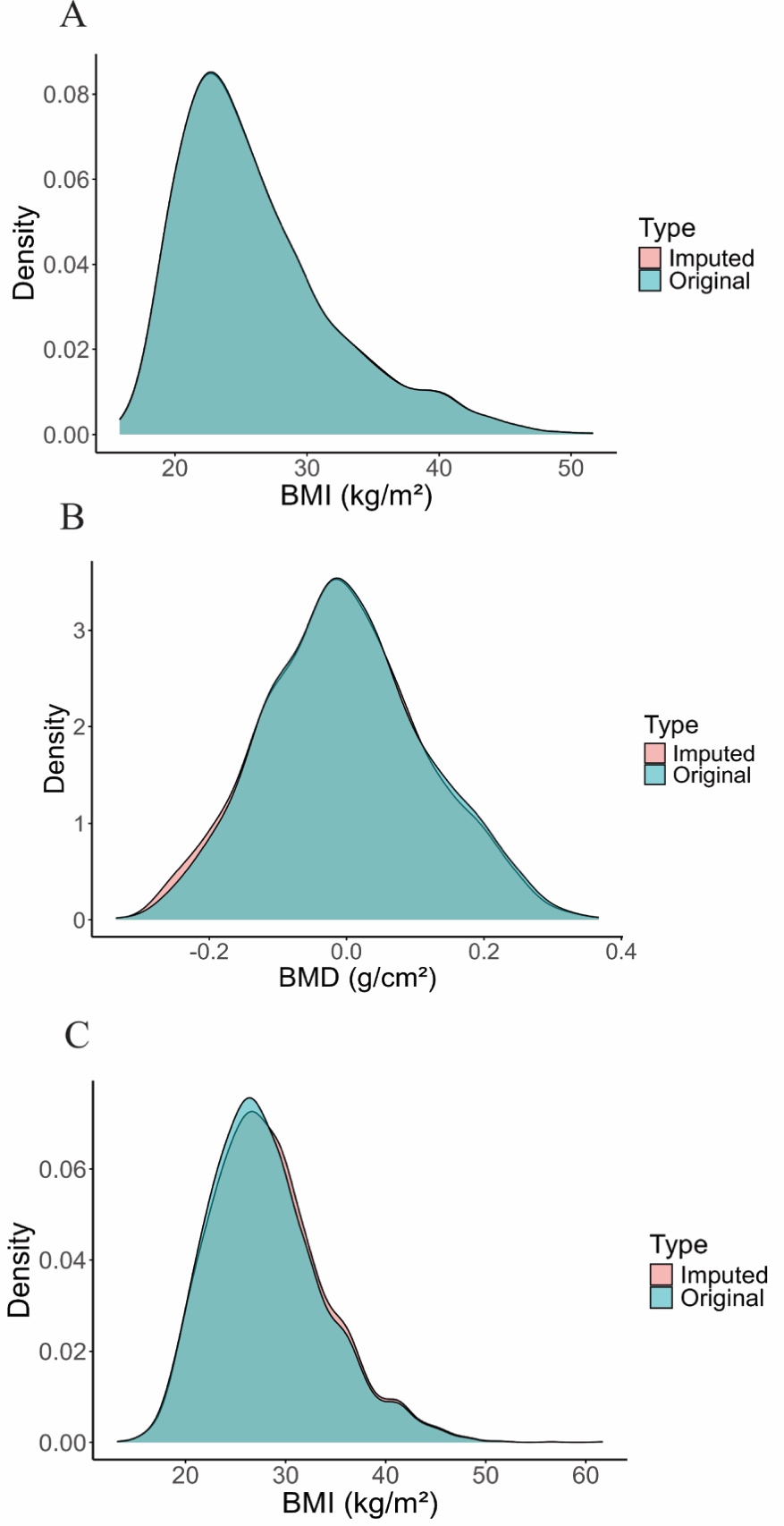

(A) BMI in the GDBF study (B) hip total BMD in the GDBF study (C) BMI in the NHANES study. The alignment of imputed and original values indicates that the imputation minimized bias and preserved the original data structure.
